# Rapid, sensitive and high-throughput screening method for detection of SARS-CoV-2 antibodies by bio layer interferometry

**DOI:** 10.1101/2020.08.15.20175851

**Authors:** Sudarshan Reddy Lokireddy, Sridhar Rao Kunchala, Ranga Pratyusha Godavarthy, Venkata Sri Krishna Kona, Laxmaiah Avula, Rakesh Kumar Mishra, Madhusudhana Rao Nalam

## Abstract

Present pandemic scenario, there exists an unmet global need for the development of a rapid and sensitive method for the detection of SARS-CoV-2 infection. The available options for identification of SARS-CoV-2 infection are detection of viral RNA by qRT-PCR, Antigen or Antibody testing by serological methods. Even though many kits available commercially but none of them are rapid, sensitive and high throughput. ‘OnCovid total antibody assay’ is a diagnostic method developed by us uses the principle of bio-layer Interferometry to detect IgM, IgA and IgG antibodies against SARS-CoV-2 antigens. This method overcomes many of the limitations normally faced in antibody detection by other methods and offers a superior platform for a rapid, sensitive and specific detection of SARS-CoV-2 infection. The test is economical, and the results can be obtained in as short as 30 seconds per test. In addition to its standalone use in early diagnosis of SARS-CoV-2, ‘OnCovid total antibody assay’ can be used to therapeutic monitoring of antiviral therapies used in clinical management and to estimate the antibody titers during convalescent plasma donation.

## Review of Literature

The novel coronavirus designated as severe acute respiratory syndrome-Coronavirus-2 (SARS-Cov-2), has caused the worst pandemic of 21^st^ century, so far, affecting almost all countries and territories around the world [1, 2]. As of August 13^th,^ 2020, 20,806,961 cases and 747,258 deaths have been reported worldwide. The current humanitarian and economic devastation caused by 2019-nCoV has triggered a race for an effective diagnostic method, which needs to be approached from all possible avenues [3].

Reverse transcriptase polymerase chain reaction (RT-PCR) is one of the best ways to detect presence of viral nucleic acids-DNA or RNA in biological samples. Detection of SARS-CoV-2 RNA can be done using nasopharyngeal swab or sputum sample, and saliva using viral gene specific probes and qRT-PCR [3–6]. Testing through qRT-PCR approaches for detection of the SARS-CoV-2 usually takes only 4-6 hours. However, the main limitation is, the typical turnaround time for shipping to referral laboratories, screening and diagnosing patients with suspected SARS-CoV-2 being > 24 h [6]. In the absence of strict quality controlled experimental conditions, the test is prone to false positives and false negatives. Further, it takes too much time, energy, and many trained personnel to run the tests.

Another diagnostic method is the antigen testing approach, wherein the presence of viral antigens in the biological samples (nasal swabs, saliva, blood etc.) is tested in a quantitative, semi-quantitative, or qualitative manner [3, 7–11]. The antigen testing requires monoclonal antibodies specific to viral antigens. An assay using a combination of different monoclonal antibodies against more than one viral antigen may result in more effective diagnosis [12]. Easy antigen testing assays which can be visualized with naked eye can be developed. However, antigen testing requires high viral count to work effectively and hence cannot be used in early detection of infection [3, 13]. Moreover, the saliva, nasal and throat swabs of asymptomatic people may not have enough antigen particles to be detected. Hence if test results are negative, it may require further confirmation by RT-PCR testing.

Yet another diagnostic approach would be to devise blood tests for antibodies against the SARS-CoV-2 virus [8–10, 12]. Systemic immune response to infection results in the production of antibodies including IgM, IgA and IgG antibodies [11, 14, 15]. ELISA tests can be used to detect specific IgM, IgA and IgG against the virus in the blood of infected patients [10, 14, 16, 17]. However, the quantity of circulating antibodies detectable by these assays will typically require > 5 days for IgM antibodies and > 10 days for IgG post infection [3, 16, 18]. However, the major limitation in the serological assays for SARS-CoV-2 is that a few infected people develop more antibodies towards viral spike glycoprotein, whereas others develop more antibodies towards nucleocapsid proteins [3, 16, 18]. Most of the available antibody detection methods rely on the spike protein, and hence leaving a void in identification of people who have antibodies against nucleocapsid protein [3, 16, 18]. Nevertheless, advanced serological testing remains one of the best methods to identify people who were recently infected, even if they are asymptomatic. The antibodies persist from very early stages of infection up to a few months, irrespective of their clinical presentation [16]. Serological tests can be used in a situation like COVID-19 pandemic for large population testing, enabling identification followed by subsequent isolation of infected and arrest potential virus spread.

Low-cost, rapid serological assays for people are far more reliable than existing thermal assays, especially useful to monitor entry/exit of individual travelers at airports and other commercial complexes. Taking into account the limitations of the existing diagnosis methods, we have developed rapid antibody detection method based on bio-layer interferometry [19–22]. Bio-layer interferometry (BLI) is a label-free optical analytical technology for measuring biomolecular interactions by analyzes the interference pattern of white light reflected from two surfaces [19–22]. The binding between a ligand immobilized on the biosensor and an analyte in solution produces an increase in optical thickness at the biosensor tip, which results in absorption properties with a wavelength shift (Δλ), which is a direct measure of the change in thickness of the biological layer [19–22]. These interactions are measured in real time, providing the ability to monitor binding specificity, rates of association and dissociation, or concentration, with high precision and accuracy [21, 22]. Any change in the number of molecules bound to the biosensor tip causes a shift in the interference pattern that can be measured in real-time.

OnCovid total antibody assay for detection of total antibodies (IgM/A/G) against SARS-CoV-2 requires ∼20 micro litres of blood or 10 micro litres of plasma sample, which can be obtained by minimally invasive finger prick method. The actual testing takes less than 30 seconds to detect the presence of SARS-CoV-2 antibodies during and after infection. In the present assay, we use virus specific ligands immobilized on the sensor and the patient plasma containing the antibodies is used as an analyte. The extent interference determined as a shift in absorbance determines the quantity of analyte present in the biological sample. The interference pattern of white light by the biosensor is taken in real time and the calculated binding rate is then correlated with standard to quantify the real amount antibody present in the given biological samples. Moreover, in OnCovid total antibody assay, biosensors can be reused multiple time (up to 50 times) which reduces cost per test very significantly.

## Results

### Material and Methods

### Chemicals and Reagents

PEI max from Polysciences, USA; 10X PBS and Ni-NTA from Bio-Rad, USA; His2 Biosensors from Fortebio, USA; Black plates 96 well plates from Greiner Bio-One, USA; Chemically defined FreeStyle-293 expression media, FreeStyle-CHO-S media, Glutamax, PenStrep, from Thermo Fisher Scientific, USA; Single used conical flask from Corning, USA; Anti-clumping agent from Irvin Scientific, USA; SARS-CoV-2 Spike S1-RBD IgG & IgM ELISA Detection Kit from Genscript, China; COVID KAWACH ELISA kit from Zydus-Cadila, India.

### Plasma or blood collections

All blood samples were collected by health professionals by abiding to human ethical approvals by Centre for Cellular and Molecular Biology (CCMB), ICMR-National Institute of Nutrition (NIN), Hyderabad, TS, India. Blood was collected from RT-PCR +Ve patients and healthy volunteers (RT-PCR −Ve) or community samples which never been tested before in K3-EDTA vacutainers according to manufacturer protocol or Finder prick collection using lancets in EDTA containing micro centrifuge tube. Plasma was separated by centrifugation at 3000 Xg for 10 minutes. Clear plasma was collected and stored at −80°C until further process.

### Cell lines and culture

FreeStyle-293 and FreeStyle-CHO-S mammalian cells from Thermo Fisher Scientific, USA were used for expression of Spike S1 subunit cloned into pcDNA3.1(+) vector. FreeStyle-293 cells and FreeStyle-CHO-S were maintained in chemically defined FreeStyle-293 expression media and FreeStyle-CHO-S media respectively.

### Transfection and Affinity purification

For transfecting FreeStyle 293/CHO-S cells, cell were grown in suspension at a density of 1×10^6^ cell/ml in growth medium and on the day of transfection, DNA: Polyethyleneimine (PEI) max (1:3 ratio) polyplexes were prepared and diluted in OptiMEM media (Invitrogen, USA) with careful mixing by pipetting (15 times) followed by incubation for 10 minutes at room temperature. Secreted recombinant S1 subunit protein was purified from filtered cell supernatant by passing through Ni-NTA resin in phosphate buffered solution (PBS) pH 7.5, 10 mM Imidazole as binding buffer and PBS pH 7.5 300 mM Imidazole as elution buffer.

### Immobilization of S1 subunit and Nucleocapsid proteins on anti-His Biosensor

The bio-layer interferometry biosensor used for this purpose has anti-His antibodies (His2 sensors) on its tip. His-tag of protein was used for binding to the biosensors’ tip by antibody-antigen affinity. C-terminal His tag containing proteins were immobilized on biosensor using inline protocol according manufactured instructions and then washed with PBS buffer. After removal of the excess unbound or loosely bound proteins from the biosensor, the biosensor is ready to use for analysis of detection and quantification of antibodies against SARS-Cov-2 in a biological plasma or blood sample.

### Recombinant SARS-CoV-2 S1 Subunit Protein (full length, 16-681 amino acids) with C-terminal His-tag

Spike S1 subunit contains a receptor-binding domain (RBD) that can specifically bind to angiotensin-converting enzyme 2 (ACE2), the receptor on target cells. Spike protein plays an important role in the induction of neutralizing-antibodies and T-cell responses, as well as protective immunity. We have cloned S1 subunit (QHD43416, Val16-Pro681) expressing clone with C-terminal His-tag in pCDNA3.1 plasmid. Secreted recombinant S1 subunit protein was purified from filtered cell supernatant by passing through Ni-NTA resin in phosphate buffered solution (PBS) pH 7.5, 10 mM Imidazole as binding buffer and PBS pH 7.5 300 mM Imidazole as elution buffer. Recombinant protein product has a calculated molecular mass of ∼75 kDa. due to the abundant glycosylation, it migrates as approximately ∼120 kDa major protein band in SDS-PAGE under DTT, β-mercaptoethanol reducing conditions.

### Recombinant SARS-CoV-2 Nucleocapsid Protein with C-terminal His-tag

We have cloned Nucleocapsid proteins (Ser2-Ala418) in pET28a(+) with C-terminal His-tag. Recombinant protein has a calculated molecular mass of ∼46 kDa. Nucleocapsid protein was purified from bacterial lysate by passing through Ni-NTA resin in phosphate buffered solution (PBS) pH 7.5, 10 mM Imidazole as binding buffer and PBS pH 7.5 300 mM Imidazole as elution buffer. Recombinant protein has a calculated molecular mass of ∼46 kDa.

### Procedure for total antibodies detection against SARS-CoV-2 in plasma using BLI method (OctetRed96)

In first step, biosensors are hydrated in PBS for 15 seconds. In second step, the hydrated biosensor is dipped for 60 seconds into a solution with His-tagged SARS-CoV-2 Spike S1 subunit and Nucleocapsid protein (Antigen) for their immobilization on the tip of the biosensor. In third step the biosensor is washed for 15 seconds in PBS to remove excess unbound protein or loosely bounded antigen. In fourth step, the ligand bound biosensor is dipped into plasma or blood samples for 90 seconds. Acquiring data of interference pattern (bio-layer interferometry sensorograms) of white light by biosensor in real-time during steps 1,2, 3 and 4, and analyzing the data using the data analysis software. The real-time data of interference pattern for all the steps of the method (steps 1-4) is provided within 180 seconds (3 minutes).

### ELISA

All ELISA experiments have been performed according the manufacturers protocol provided with kit. The difference between two ELISA kits are, the Genscript ELISA kit use RBD domain as antigen whereas COVID KAVACH ELISA kit uses gamma irradiated whole SARS-CoV-2 virus as antigen [23].

### Statistical analysis

Statistical Analysis was performed using Student’s t tests, One-way ANOVA post hoc Bonferroni’s comparison, Receiver operating characteristic (ROC) curve analysis for sensitivity and specificity of tests and Pearson r performed for Correlation. All values are expressed as means ± SD.

### Bio-layer interferometry (BLI) method for detection on total antibodies against SARS-CoV-2 virus (ethical clearance needs a mention)

“Oncovid total antibody assay” is a diagnostic assay developed by us is a one-step antigen capture format on a Biosensor, intended for the qualitative and quantitative detection of total antibodies including IgM/A/G to SARS-CoV-2 in human serum and plasma using the principle of BLI technology. The “OnCovid total antibody assay” assay is intended for use as an aid in identifying individuals with an adaptive immune response to SARS-CoV-2, indicating recent or prior infection. All blood/plasma samples were collected from COVID-19 patients or healthy volunteers or community by health professionals by abiding to human ethical approvals by CSIR-Centre for Cellular and Molecular Biology (CCMB), ICMR-National Institute of Nutrition (NIN), Hyderabad, TS, India.

A typical pattern to detect total antibodies against SARS-CoV-2 virus in plasma using the BLI method includes four steps. The biosensors get hydrated in 1X PBS followed by antigen capture and washing to remove the excess or loosely bounded antigen in the second and third steps respectively (Figure-S1A). Finally, the antigen coated biosensor shall be dipped into 20 times diluted plasma or 10 times diluted blood samples for analyte analysis (Figure-1A). Acquiring data of interference pattern (biolayer interferometry sensorograms) of white light by biosensor in real-time during steps 1, 2, 3 and 4, and analyzing the data can be monitored (Figure-1B).

**Figure-1.**
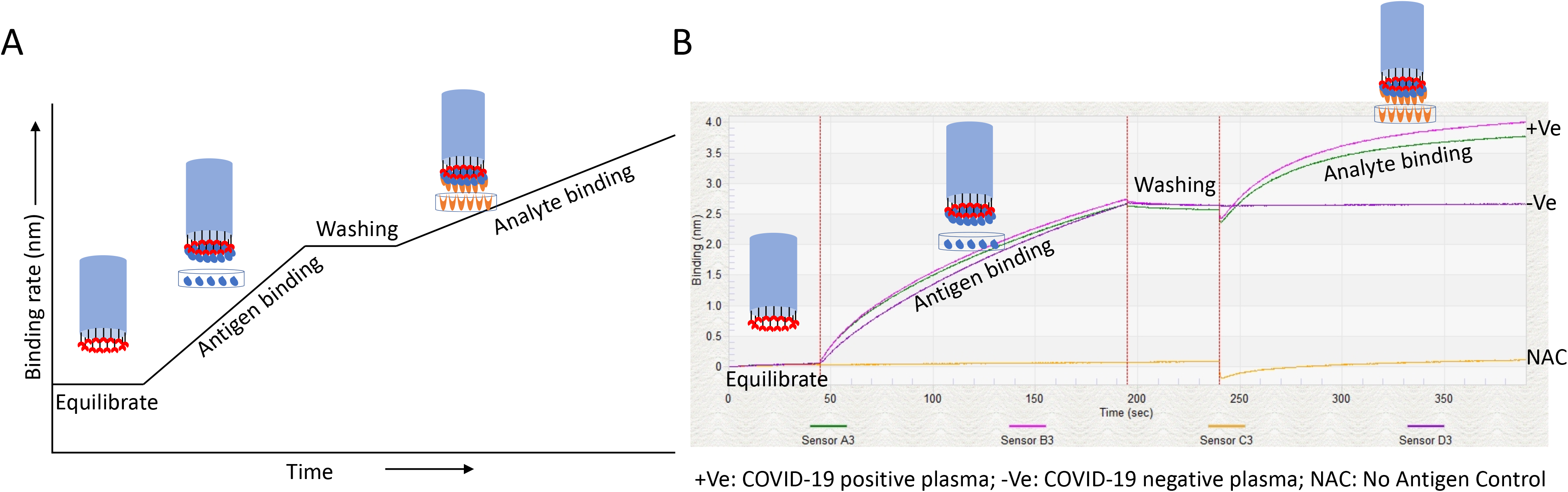
**A)** Graph represents typical pattern to detect total antibodies against SARS-CoV-2 virus in plasma. In first step, biosensor shall be hydrated in 1X PBS. In second step, after hydration the biosensor shall be dipped into His-tagged SARS-CoV-2 Spike and Nucleocapsid protein(Antigen) for immobilizing on the tip of the biosensor. In third step, after antigen binding, the biosensor shall be washed in 1X PBS to remove excess unbound protein or loosely bounded antigen. In fourth step, after washing of ligand bound biosensor shall dipped into plasma or blood samples. Acquiring data of interference pattern (biolayer interferometry sensorograms) of white light by biosensor in real-time during steps 1, 2, 3 and 4. In this method Biosensors can be reuse up to 50 times. **B)** Binding curve represents plasma samples collected from patients which are diagnosed with RT-PCR (+Ve) and RT-PCR negative (−Ve). No antigen control (NAC) have been used as internal control and background correction. Plasma samples used a NAC is a RT-PCR positive plasma. Plasma diluted 1 to 20. 500ng/ml of Spike S1 proteins has been used as antigen. We called this method as OnCovid total antibody assay.

To test this idea, we had performed an experiment using Spike S1 subunit of SARS-CoV-2 as antigen [11] against to COVID-19 RT-PCR positive (+Ve) and negative (−Ve) plasma samples at 1 to 10 dilution (Figure-1B and Figure-S2A). The positive (+Ve) plasma for SARS-CoV-2 infection shows a sharp raised sensogram but not in negative (−Ve) plasma sample (Figure-1B). However, when positive plasma for SARS-CoV-2 incubated with biosensors without antigen (No antigen control (NAC)) there wasn’t any raise in sensogram, indicate that the raise in sensogram with seropositive plasma when antigen present is indeed positive for antibodies against SARS-CoV-2 (Figure-1B).

### Identification of suitable antigen and dilution factor of plasma to detect total antibodies against SARS-CoV-2 infection

To increase the sensitivity of detection of total antibodies against SARS-CoV-2, we have tested another abundant and highly immunogenic protein-the Nucleocapsid phosphoprotein [3, 16]. In RT-PCR based diagnosis, the nucleocapsid protein of SARS-CoV-2 is often used as a marker. A recombinant C-terminal His-tag containing Nucleocapsid has been expressed in E. coli and affinity purified was used to coat the biosensors. (Figure-S2B). To our surprise, we have found that there were significantly higher antibodies (IgM/A/G) against the Nucleocapsid proteins compared to Spike S1 subunit in SARS-CoV-2 positive plasma sample (Figure-2A).

**Figure-2.**
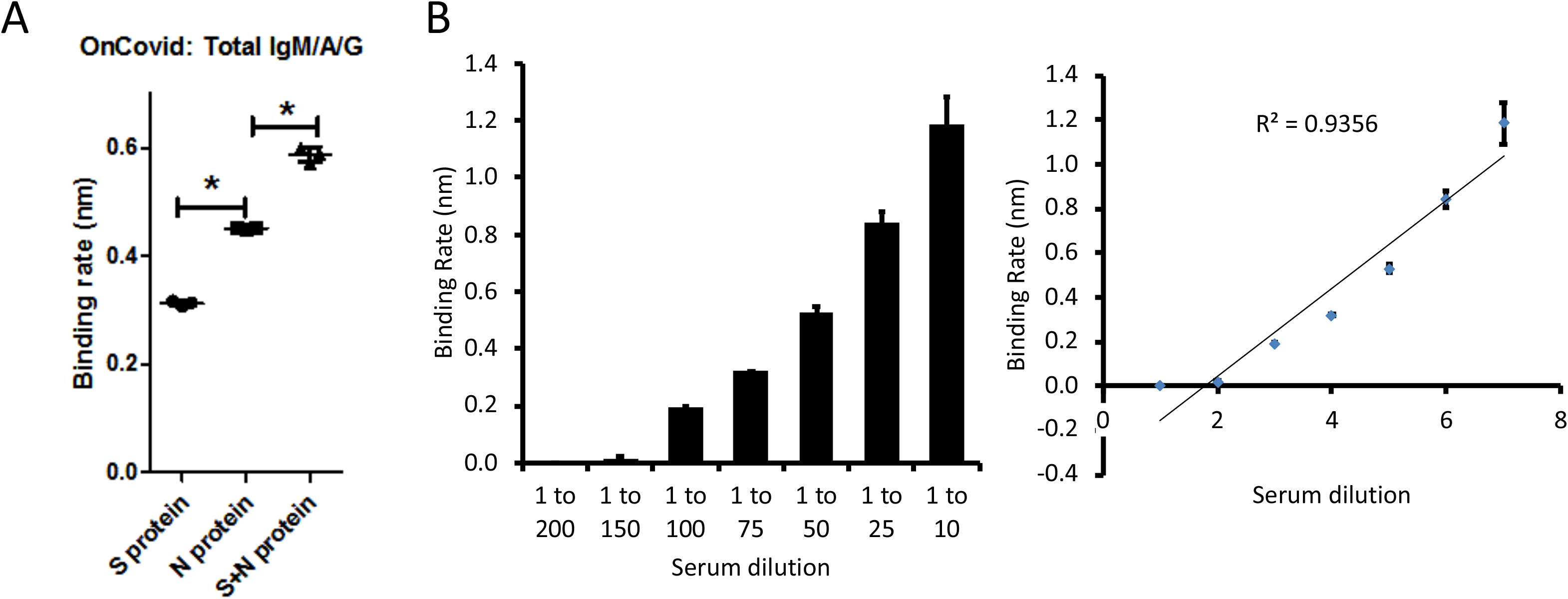
**A)** Graph represents binding rate constant for each said protein or together against plasma samples collected from RT-PCR positive patients. Three independent samples were analysed which have similar binding rate. A molar ratio of 1:1 has been used for Spike S1 and Nucleocapsid proteins in all studies. Binding rate has been calculated based on standard curve equation, Linear Point to Point and binding rate equation using R equilibrium using Data Analysis Software provided by ForteBio. Error bar represents standard deviation from three independent samples. * P < 0.0001. **B)** Bar graph and line represents the binding rate of different dilutions of plasma from RT-PCR positive patient. During data analysis reference was subtracted prior to fitting performed using the ForteBio data analysis software. Based on dilution and no antigen control, 0.099 nm considered as negative and 0.1 nm considered as positive. Error bar represents standard deviation from two independent samples.

Moreover, when an equimolar ratio of Spike S1 and Nucleocapsid proteins were used for immobilization on the sensor, we noticed significant increase in binding rate of antibodies in SARS-CoV-2 positive plasma compared to their standalone use (Figure-2A). However, we couldn’t observe doubling of the binding rate when two proteins (Spike S1 subunit and Nucleocapsid protein) combined, this could be due to saturation protein binding sites on the biosensors (Figure-2A).

Next, we also tested different dilutions of plasma on both antigens Spike S1 and Nucleocapsid to find sensitivity and specificity of OnCovid total antibody assay. Two independent SARS-CoV-2 positive plasma samples with identical binding rates were diluted up to 200 times with PBS and binding rate has been calculated using Data Analysis software provided by ForteBio. Though the total antibodies have been detected up to a 100^th^ dilution with binding rates close to 0.20 (Figure-2B), we observed a linear binding rate up to 1 in 10 dilution having R^2^ value of 0.9356 (Figure-2B). Based on these results we had used 1 to 20 dilution of plasma in all further experiments. A binding rate with a value of 0.10 nm considered as positive and 0.099 nm considered as negative.

### “OnCovid total antibody assay” is more sensitive and specific compared to ELISA

To test specificity and sensitivity of ‘OnCovid total antibody assay’, we performed the assay with plasma samples collected before (plasma collected month of November 2019, (40 samples)) and after COVID-19 pandemic situation (plasma collected from SARS-CoV-2 patients during April 2020 (3 samples)). In all 40 samples collected before COVID-19 pandemic didn’t shows antibodies against Spike S1 subunit and Nucleocapsid proteins(Figure-S3). However, plasma samples collected from SARS-CoV-2 infected patients show significant quantities of antibodies against Spike S1 and Nucleocapsid proteins (Figure-S3). Moreover, Receiver Operating Characteristic (ROC) curve analysis for detection of antibodies against to SARS-CoV-2 by OnCovid total antibody assay shows 100% sensitivity and specificity (Figure-S3).

Next, we compared OnCovid total antibody assay with commercially available ELISA kit (SARS-CoV-2 Spike S1-RBD IgG & IgM ELISA Detection Kit from Genscript, China). A total of 17 RT-PCR −Ve and 31 RT-PCR +Ve were analysed for antibodies by ‘OnCovid total antibody assay and Spike S1-RBD IgG & IgM ELISA. ‘Oncovid total antibody assay’ detected antibodies in all the 31 RT-PCR +Ve plasma samples. ‘OnCovid total antibody assay’ couldn’t detect any antibodies in RT-PCR −Ve plasma (Figure-3A). GenScript ELISA kit however, detected IgM in 24 and IgG in 29 RT-PCR +ve samples, (Figure-3B and 3C). GenScript assay also showed positivity of in one sample for IgM and in 4 samples for IgG in RT-PCR −Ve plasma (Figure-3B and 3C). Further, we had performed ROC curve analysis on this data found that 100% specificity and sensitivity with ‘Oncovid total antibody assay’(Figure-3A). GenScript kit shows ROC curve area only 0.9023 for total IgG and confidence interval in between 0.8100 to 0.9945 and ROC curve area shows only 0.9070 for total IgM and confidence interval in between 0.8243 to 0.9898 (Figure-3B and 3C).

**Figure-3.**
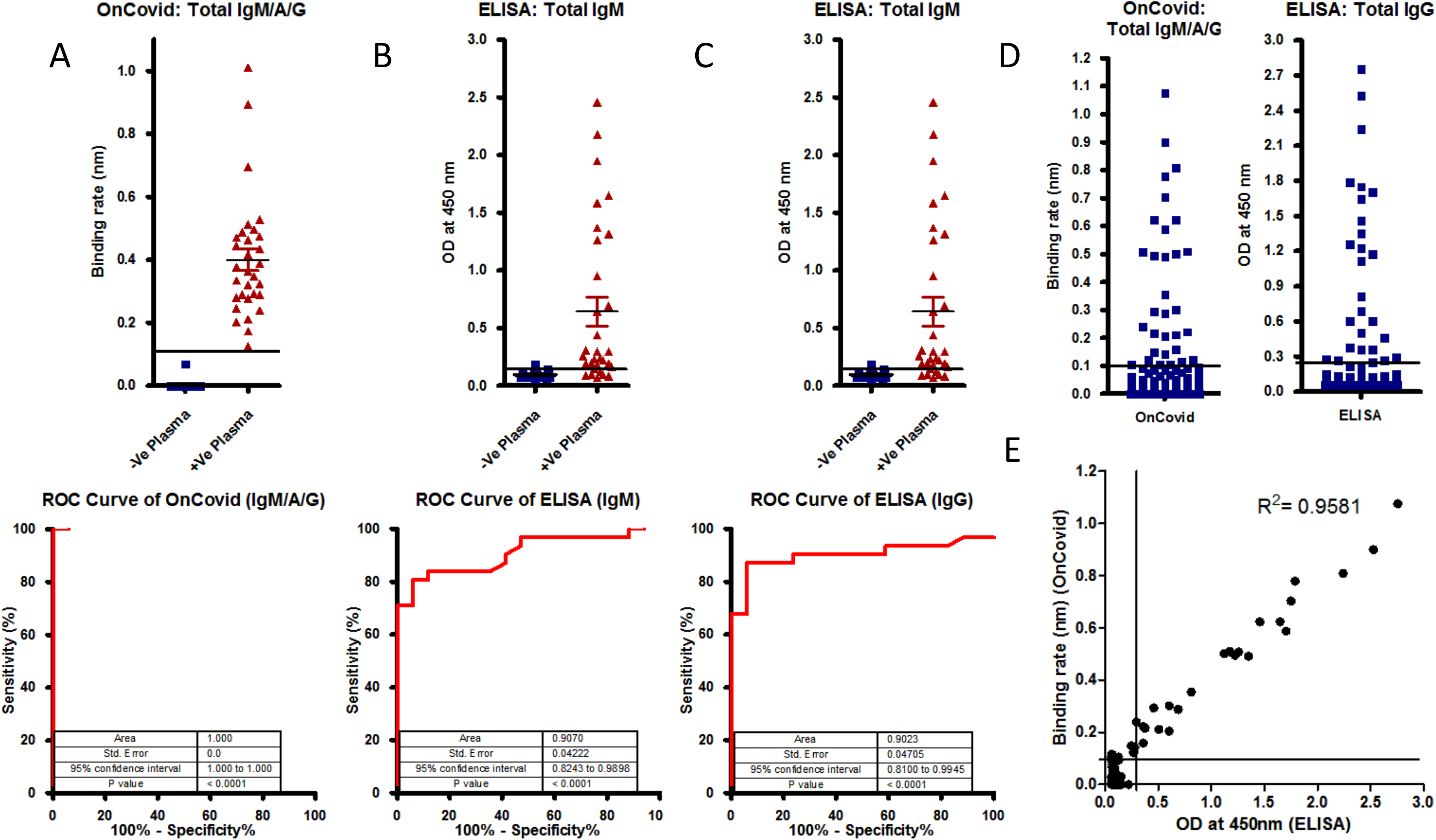
**A)**, **B)** and **C)** A total of 17 RT-PCR −Ve and 31 RT-PCR +Ve samples were tested using commercial ELISA kit (GenScript SARS-CoV-2 Spike S1-RBD IgG & IgM ELISA Detection Kit) for both IgG and IgM antibodies detection in SARS-CoV-2 infected patients and healthy controls. OnCovid total Ab assay ROC area shows 1.000, confidence interval in between 1.000 to 1.000, and p values < 0.0001, whereas for total IgG shows only 0.9023 and confidence interval in between 0.8100 to 0.9945 and for total IgM shows only 0.9070 and confidence interval in between 0.8243 to 0.9898. Line indicate cut off point. **D)** A total of 486 samples which never tested for SARS-CoV-2 were analyzed by OnCovid total antibodies assay and ELISA kit (COVID KAVACH ELISA) developed by National Institute of Virology, India using killed SARS-CoV-2 viral particles. A total 30 plasma samples were shown positive by ELISA kit and 32 plasma samples by OnCovid total antibody assay. Line indicate cut off point. **E)** Both ELISA (COVID KAVACH ELISA) and OnCovid total antibody assay shown a good correlation at R^2^ = 0.9581, Person r = 0.9788 and p value < 0.0001.

We have also compared ‘OnCovid total antibody assay’ with COVID KAWACH ELISA [23] in large set plasma samples (486) collected from community where individual never been diagnosed for COVID-19 infection. In COVID KAWACH ELISA kit, gamma irradiated whole viral particle is used as antigen. Based on previous report, this kit known to have more than 95% specificity and sensitivity against SARS-CoV-2 [23]. COVID KAWACH ELISA kit detected only 30 samples as antibody positive against SARS-CoV-2 whereas ‘OnCovid total antibody assay’ detected 32 samples as positive (Figure-3D). We found more than 95 % correlation in detection with ‘OnCovid total’ and COVID KAWACH ELISA kit (Figure-3D and 3E). In total, 4 positive samples analysed by OnCovid total antibodies assay differed with COVID KAWACH ELISA kit (Figure-3E). This difference could be due to the fact that COVID KAWACH ELISA kit detects only anti-IgG and not anti-IgM antibodies against SARS-CoV-2. Detection of IgM antibodies is of paramount importance to diagnose infection in early stages. The ‘OnCovid total antibody assay’ kit shows greater sensitivity and specificity in detection of IgM, IgA and IgG antibodies against SARS-CoV-2 infection.

### ‘OnCovid total antibody assay’ on community surveillance

Using OnCovid total antibodies assay, we tested a total of 1610 plasma samples collected from different communities and hospitals. Out of 1610, only 123 samples were tested for RT-PCR +Ve and rest of the samples never diagnosed for COVID-19. Out of 1487 samples which never diagnosed for COVID-19, 81 samples have shown significant levels of antibodies against SARS-COV-2 (Figure-4A and 4B). Moreover, in 4 samples tested for RT-PCR +Ve didn’t show antibody against SARS-CoV-2 beyond the threshold levels. (Figure-4A). This could be due to early detection of viral infection by RT-PCR. Furthermore, in overall specificity and sensitivity by ROC curve analysis demonstrated that curve area of 0.9818, at 95% confidence interval in between 0.9663 to 0.9973 and p < 0.0001. In this large set of data demonstrated that ‘OnCovid total antibody assay’ is quite sensitive and specific to SARS-CoV-2 infection.

**Figure-4.**
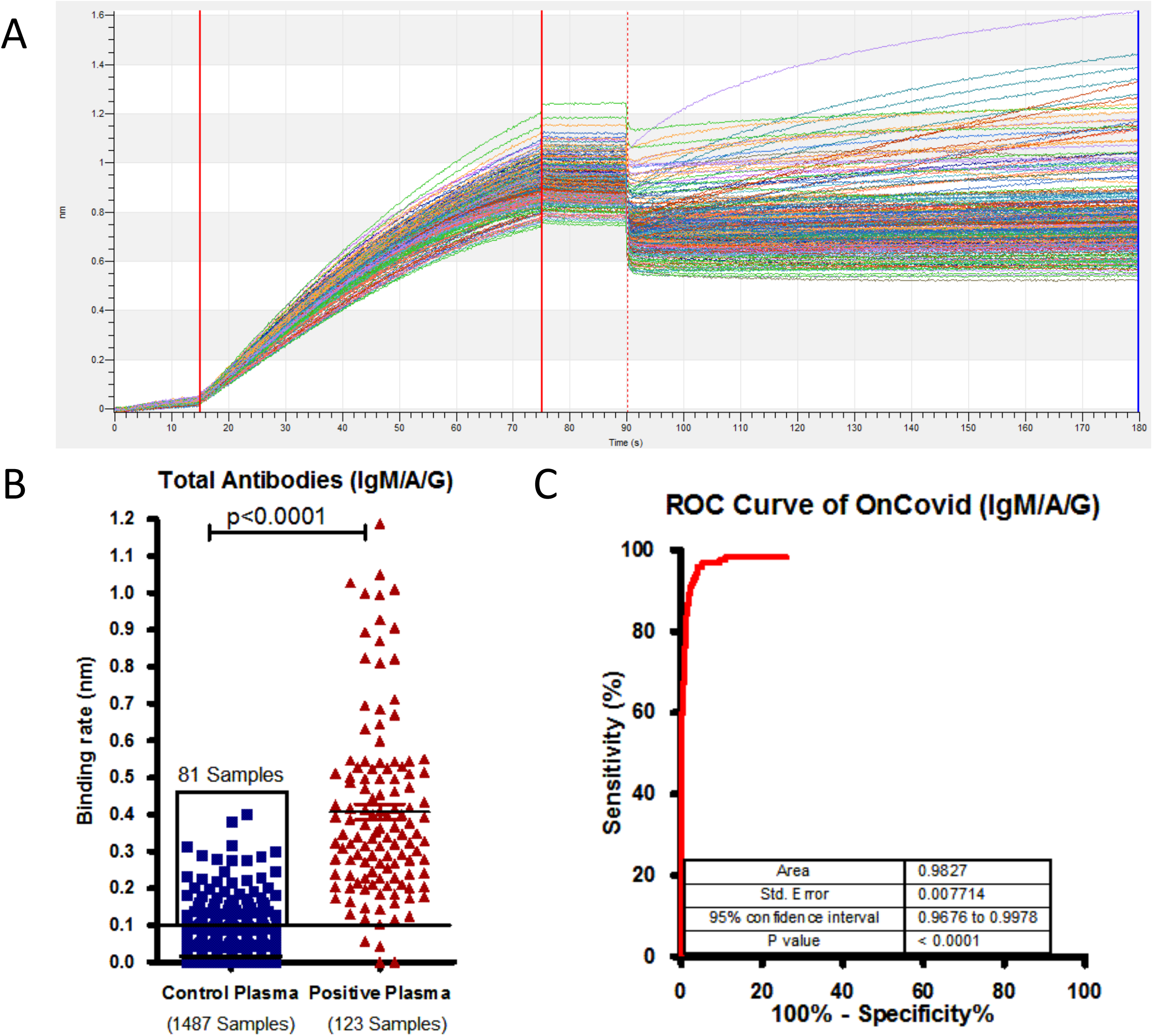
**A)** Binding curve represents detection of total antibodies against SARS-CoV-2 in plasma samples collected from a community who were never being diagnosed for SARS-CoV-2. **B)** Binding rate have been calculated on a total of 1487 plasma samples collected from a community who were never diagnosed for SARS-CoV-2 and 123 plasma collected from diagnosed for SARS-CoV-2 using RT-PCR. Found that 81 samples were shows binding rate above 0.1nm out 1487 control sample tested. Line indicate cut off point. **C)** Receiver operating characteristic (ROC) curve analysis showing sensitivity versus specificity for discrimination of RT-PCR positive and control plasma samples which were never tested for SARS-CoV-2 (ROC area under the curve: 0.9818, 95% confidence interval in between 0.9663 to 0.9973 and p values < 0.0001).

### Detection total antibodies in the blood using OnCovid total antibody assay

To reduce the time for preparing of samples, we tested direct blood samples (20µl) of 2 SARS-CoV-2 RT-PCR +Ve and 5 RT-PCR −Ve samples by comparing with respective to plasma for binding rate. Bio-layered interferometry sensogram shows perfect correlation in total antibodies present in blood and (Figure-5A, 5B and 5C). Moreover, the correlation analysis also shows good correlation between blood and plasma data at R2 = 0.9961 and pearson r = 9980 and p = 0.0001. The data clearly conclude that OnCovid total antibody assay can performed in whole blood. However, care should be taken to prevent the lysis of RBCs while collecting the sample.

**Figure-5.**
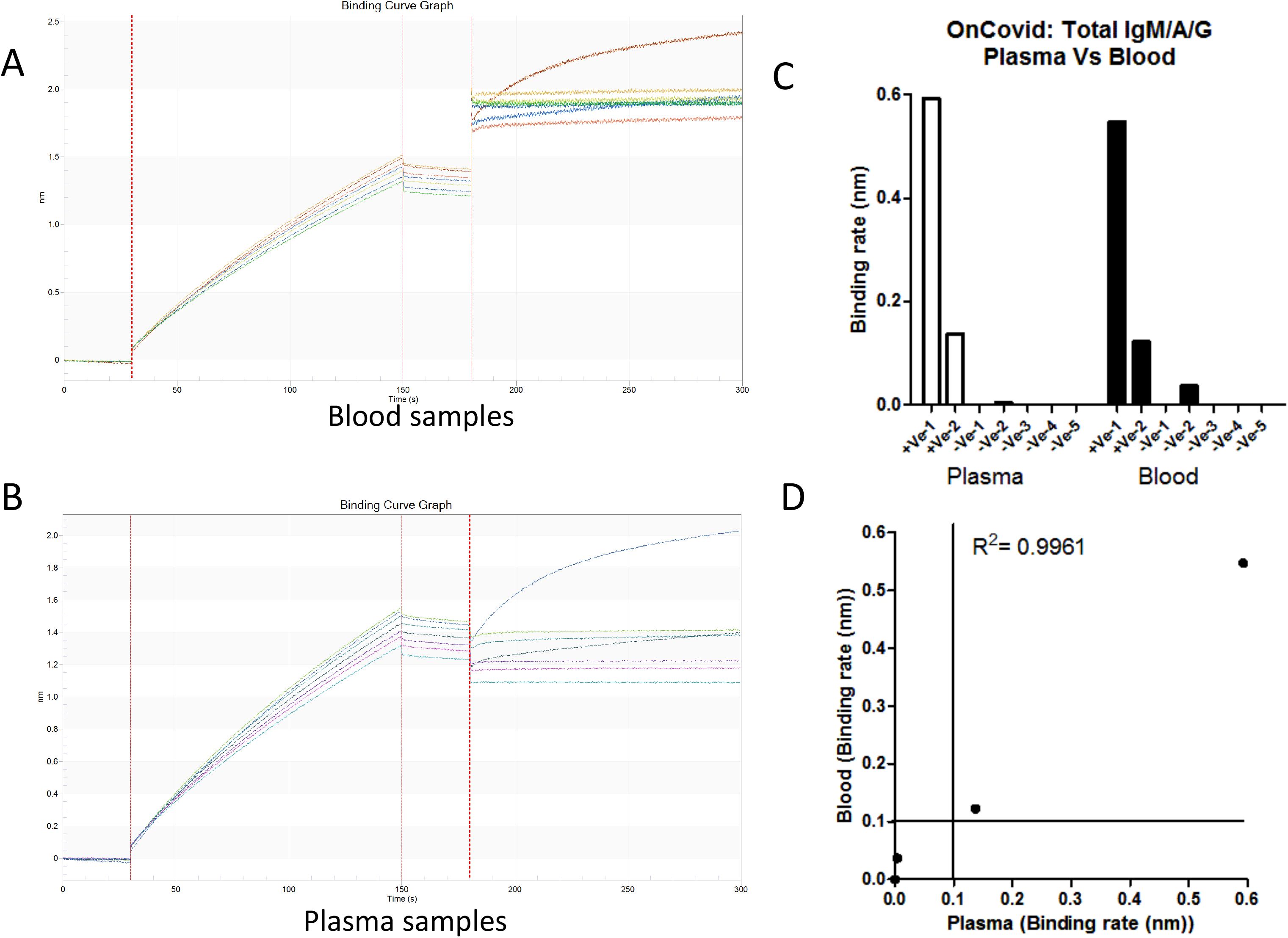
**A)** and **B)** Binding curve represents detection of total antibodies against SARS-CoV-2 in 20 µl blood (A) and 10µl plasma (B) samples collected from patient diagnosed for SARS-CoV-2 (2 seropositive) and healthy volunteer (5 seronegative). **C)** Binding rate have been calculated on both blood and plasma samples collected RT-PCR positive patients and healthy controls. **D)** Both Blood and Plasma shows a correlation at R^2^ = 0.9961, Person r = 0.9980 and p value < 0.0001. Line indicate cut off point.

## Discussion

The basic structure of SARS-CoV-2 is similar to any other coronavirus comprising of spike glycoprotein (S), membrane protein (M), envelope protein and nucleocapsid protein (N). SARS-CoV-2 spikes are composed of trimers of spike glycoprotein [17]. The spike glycoprotein is the main antigenic component in the SARS-CoV-2 and an important target of host defense system for producing neutralizing antibodies [24]. Hence, the spike glycoprotein is a common target for vaccine and therapeutic development [24]. The nucleocapsid protein is the most abundant protein in SARS-CoV-2 and is required for coronavirus RNA synthesis and has RNA chaperone activity that may be involved in template switch [13]. It is a highly conserved and immunogenic phosphoprotein. The nucleocapsid protein of SARS-CoV-2 is often used as a marker in diagnostic assays [13]. Although reliable experimental approaches and detection technologies for diagnosis of latent infection or active COVID-19, including RT-PCR, ELISA, and LAMP exist, but they are either more expensive or time-consuming or not high throughput [3, 9, 10, 13]. Global comparative studies of existing technologies, it is our conclusion that rapid development and high throughput screening of antibodies against SARS CoV-2 virus by BLI method would be rapid and can be detected in less than a minute and under a dollar per test. OnCovid total antibody assay based on BLI method can be used for detecting and quantifying of total antibodies (IgM/A/G) in serum or plasma or blood in less than a minute per test. BLI method is very high sensitivity and specificity for detection of COVID-19 compared to the existing technologies since this approach will be using a pool of recombinant protein to capture anti-2019-nCoV IgM/A/G in serum during and after SARS CoV-2 infection symptoms appears in community studies or vaccine development or in plasma therapy.

Data presented here clearly demonstrated OnCovid total antibody assay using BLI method is sensitive and specific in detection of total antibodies against SARS-CoV-2. The OnCovid total antibody assay is simple to perform and also have less time for sample process compared any other ELISA based diagnosis. OnCovid total antibody assay can be performed in 96 or 384 formats. Moreover, the assay is rapid and quantitative for detection of total antibodies with real-time data monitoring full results in less than half minute per sample in 96 well format and 15 seconds for sample in 384 well format.

In addition to shorter duration of assay time, OnCovid total antibody assay has several technical advantage over traditional ELISA, CLIA and IFA [3, 9, 10, 13]; first, OnCovid total antibody assay is a label free and gives information on the accurate quantities of antibodies unlike other ELISA systems that gives ‘relative’ values; secondly, there are no ‘storage related’ changes as the complete assay is done on a real time; third, this method does not require additional steps such as washing, incubation, substrate addition etc.; fourth, direct blood can be used to detect total antibodies; fifth, high throughput with 8 and 16 biosensors at a time and advanced equipment such as Octet HTX can measure 96 biosensors at a time; sixth, less technical team required; seventh, data variability is less; eighth, finger prick blood sufficient to do the assay and last but not least biosensors can be reused up to 50 times. Besides of these advantages, we do understand that to perform OnCovid total antibody assay required high capex instrument which is one-time investment.

## Conclusion

OnCovid total antibody assay uses BLI platform to detect antibodies against SARS-CoV-2 using Spike S1 subunit and Nucleocapsid protein. Both proteins have shown good correlation with whole virus particle ELISA kit and Spike RBD kit. Our OnCovid total antibody assay based BLI method could be potential game changer since it has shown high sensitivity and specificity for detection of COVID-19 compared to the existing technologies. Moreover, this approach will be using a pool of recombinant protein to capture anti-2019-nCoV IgM/A/G in serum/plasma/blood during and after SARS CoV-2 infection. This assay method could test in community studies or vaccine development or in convalescent plasma therapy.

## Data Availability

All raw data files shall be available upon request.

## Acknowledgements

Authors are grateful for the support from volunteers who donated blood to present study.

We like to Ramaswamy P and Chennakesavulu T from Oncosimis Biotech for helping in cell culture and protein expression in E.coli. This technology has been selected by C-CAMP COVID-19 innovations deployment accelerator (C-CIDA) and partially funded and supported by CSIR-CCMB, Hyderabad under Deploying COVID-19 Innovation. Oncosimis Biotech Private Limited also grateful for DBT, BIRAC for providing funding for AcceTT® platform for increased expression of recombinant protein in mammalian system.

## Authors contribution

SRL, SRK, MRN and RKM conceived the study. SRL, SRK, RPG, and VSKK expressed Spike S1 in FreeStyle 293 cells and purified proteins. MRN and RKM collected blood samples and ethical approval. MRN provided Nucleocapsid protein. LA collected community samples and performed COVID KAWACH ELISA. SRL, SRK, MRN and RKM analyzed the data. SRL and MRN prepared the figures. SRL, SRK and MRN wrote the manuscript. All authors edited the manuscript and approved.

---

**Figure-S1.**
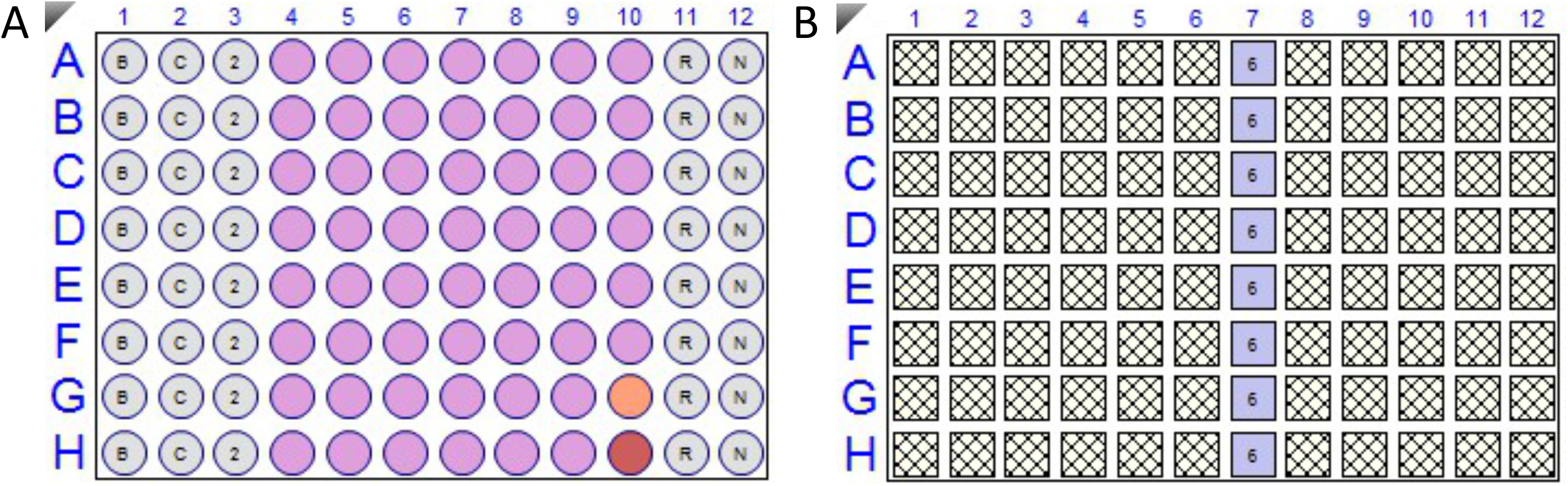
**A)** A typical 96 well plate arrange for OnCovid total antibody assay. R represents Regeneration buffer (10mM Glycine pH 2.0); N represent Neutralization buffer (PBS); B represents Buffer-1 (PBS), C represents Capture antigen (Spike S1 and Nucleocapsid proteins), Pink wells for samples (Plasma or blood), Orange for control and Red for Reference for subtraction. **B)** A typical Biosensor plate for 8 channel OctetRed 96 or any similar kind of machine.

**Figure-S2.**
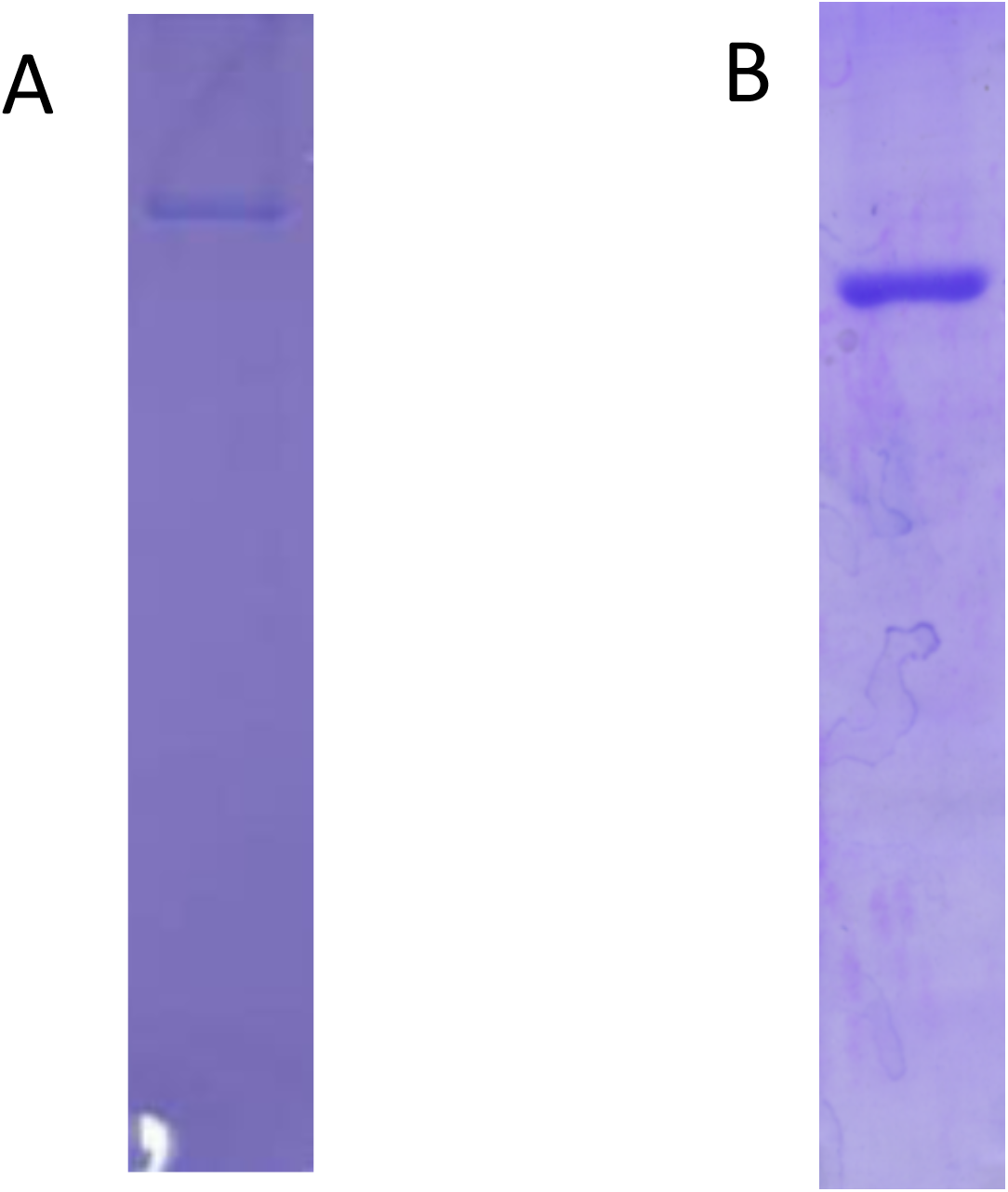
**A)** & **B)** A total of 5 ug was loaded after affinity purification of S1 protein (A) and N protein (B) using Ni-NTA resin from FreeStyle 293 cells supernatant and Bacterial lysate in 8% and 12% SDS-PAGE gel respectively.

**Figure-S3.**
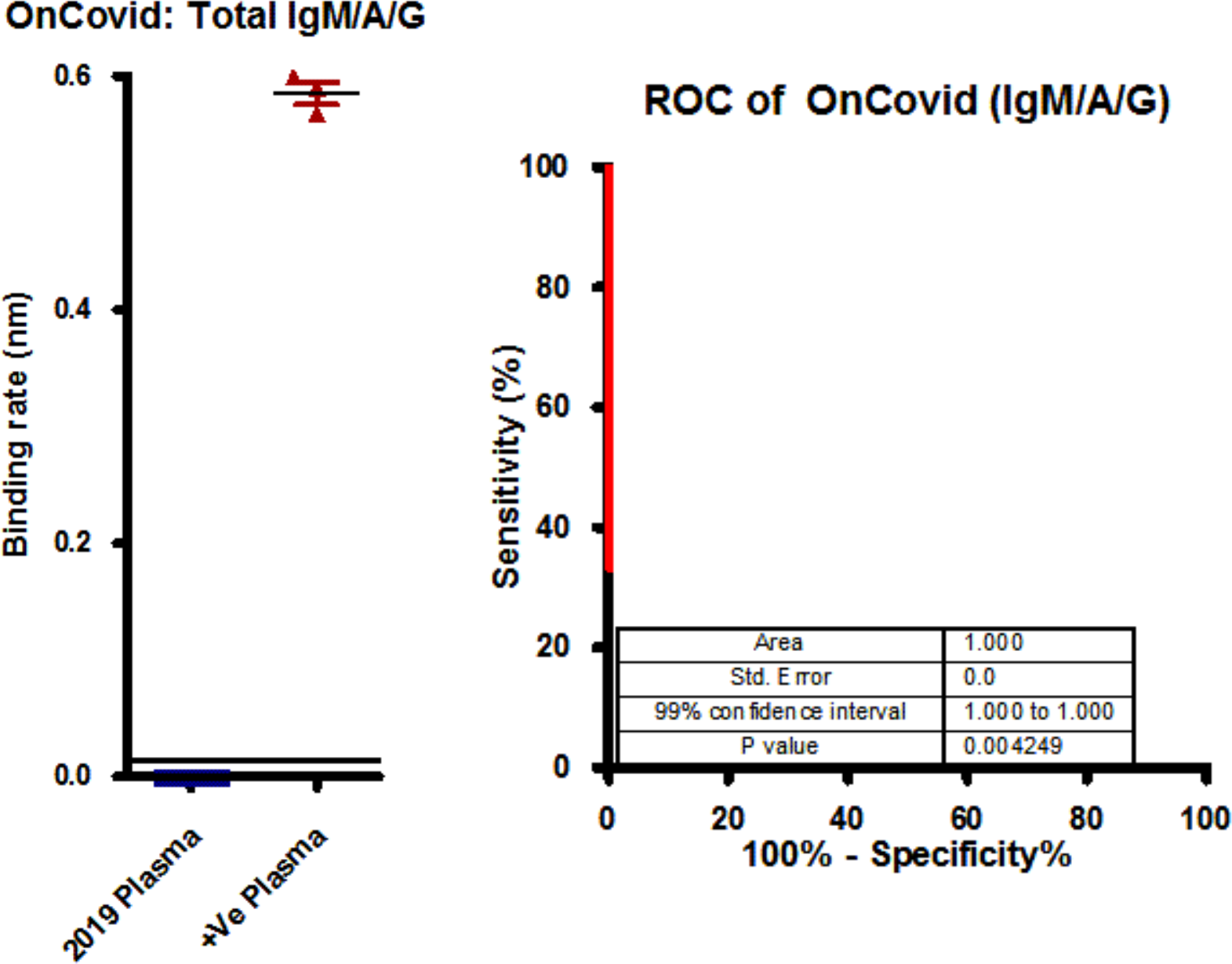
Graph represents the binding rate (Left graph) of plasma samples (40) collected before COVID-19 pandemic (Nov 2019) and after COVID-19 pandemic (3) against S1 subunit and Nucleocapsid antigen. Receiver operating characteristic curve (ROC) (Right graph) analysis showing sensitivity versus specificity for discrimination of RT-PCR positive and plasma samples collected before COVID-19 pandemic (ROC area under the curve: 1.000, 95% confidence interval in between 1.000 to 1.000 and p values 0.004249).

